# Reopen or redistribute? - Modeling years of life lost due to Covid-19, socioeconomic status, and non-pharmaceutical interventions

**DOI:** 10.1101/2021.04.23.21256005

**Authors:** Jari John

## Abstract

Research in the current pandemic has put a sharp focus on the health burden of Covid-19, thereby largely neglecting the cost to life from the socioeconomic consequences of its containment. The paper develops a model for assessing their proportionality. It compares the years of life lost (YLL) due to Covid-19 and the socioeconomic consequences of its containment. The model reconciles the normative life table approach with *de facto* socioeconomic realities by correcting YLL estimates for socioeconomic differences in life expectancy. It thereby aims to improve on the attribution of YLL due to immediate and fundamental sources of inequalities in life expectancy. The application of the approach to the pandemic suggests that the socioeconomic consequences of containment measures potentially come with a much higher life tag than the disease itself and therefore need urgent attention, especially in poorer and more unequal societies. Avoiding 3 million additional cases of extreme poverty may come with a similar life tag as protecting 1 million people from dying from Covid-19.

## 1. Introduction

The Covid-19 pandemic has caused severe suffering. More than 3 million people have lost their lives to Covid-19 with estimates projecting up to 5 million deaths by August 2021.^1^ To contain the spread of the virus, governments around the world mainly relied on non-pharmaceutical interventions (NPI). These came with a heavy socioeconomic burden, however, especially for the poor. According to ILO estimates, in 2020 8.8% of all working hours were lost (the equivalent to 255 million full time jobs).^2^ Remittances to poorer countries declined dramatically. The UNDP warns that up to 1.6 billion people could lose up to half of their income.^3^ Extreme poverty could increase by 100-150 million people (or twice as many under the less strict poverty line of $3,20 per day).^4,5^ Prolonged school closures that temporarily affected up to 1.5 billion students will depress long-term economic recovery.^6^ Whether the long-term socioeconomic harm outweighs the benefit to protect health in the short-term is therefore a key question in the pandemic.

Governments justify the use of NPIs by referring to their proportionality. Three component parts define the proportionality of NPIs. The first two concern their efficacy, i.e. the suitability and necessity of particular measures. They largely belong to the realm of epidemiologists and virologists and are beyond the scope of the paper.^7–9^ The third meaning concerns the proportionality of NPIs in the narrower sense. It asks whether they are reasonable given the competing interest of parties. In the pandemic at least two problems complicate such an assessment, however. Subjective risk perceptions tend to suffer from significant distortions, rendering public citizen assessments of proportionality of limited reliability.^10–12^ More importantly, no common measure exists to compare the immediate health threat from Covid-19 to the mostly indirect long-term socioeconomic harm from NPIs. Such efforts are further complicated by important moral and legal concerns against weighing lives against lives in the pandemic.^13^ Furthermore, rather than being a great equalizer the pandemic has exacerbated existing inequalities, leaving one and the same people most exposed to health and socioeconomic risks. Any such comparison must therefore primarily aim at gauging the extent of proportional socioeconomic compensation and raising the burden of proof for suitability and necessity of NPIs, especially in the context of resource scarcity and gravity of consequences (e.g. extreme poverty). After all, people can be lifted form poverty but not be resurrected from the dead. Therefore, the question of proportionality can only be meaningfully answered in the future.

Against this background the paper introduces a model to compare the damage to life from Covid-19 and the socioeconomic consequences of NPIs. Starting point of the considerations is that both acute infectious diseases such as Covid-19 as well as a low socioeconomic status (SES) may shorten an individual’s life expectancy. Accordingly, it is possibly to assess the damage due to Covid-19 and NPIs in years of life lost (YLL). The approach complements but is distinct to other perspectives on the pandemic such as the burden of disease and value of life. The model rather contributes to the discourse on the relationship between health inequality and social justice.^14,15^ The paper targets some of the key conceptual difficulties when attributing YLL to individual causes. Because any such assessment can only be plausible estimates at best, the efforts at quantifying the model are primarily for purposes of illustration and need to be complemented with context-specific data to be able to inform policy.

## 2. Methods: Measuring YLL due to NPIs and Covid-19

The model starts from the basic assumption that proportionality can be expressed as a correspondence of YLL due to Covid-19 and the socioeconomic damage from the NPIs.

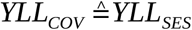

YLL refer to the gap between the age of death and the age to which a person could have lived. No single methodology for estimating YLL exists, but it is common practice to use life tables that either assume an ideal life expectancy in a counterfactual disease and poverty free, egalitarian world (or society) or draw on hazard ratios within the age bracket of the birth cohort.^16,17^ As a result life tables rather state an aspiration than provide information about the actual number of years an individual would have lived in the absence of a specific cause of death. While there is nothing wrong about such a normative approach, it entails problems of correctly attributing YLL to individual causes. The question therefore is to what extent these YLL can be attributed to the immediate cause of death or in fact reflect more fundamental socioeconomic differences in life expectancy.

International differences in average life expectancy amount to about 30 years between the poorest and the richest countries.^18^ Accordingly, the WHO’s international life tables state an average of more than 50 YLL for a death in a low income country compared with less than 20 YLL in a high income country.^19^ Those numbers roughly halve when correcting for the average life expectancy in the income group. National life tables such as those of UN’s World Population Prospects take into account international differences in life expectancy but fall short of accounting for systematic differences in life expectancy among socioeconomic groups within a society. By common ways of measurement socioeconomic differences in life expectancy in high income countries usually amount to 5-10 years between groups with a low and high SES.^20^ These differences may reach up to 15 or 20 years in poorer and more unequal societies and when using more fine-grained measures.^21,22^ The common tripartite division in low, mid, and high SES means that a person that may fall into a lower SES group, for example, as a consequence of losing around 40% of their income due to unemployment or a transition from full-to part-time work, or a forgone 3-4 year higher education block (e.g. secondary or tertiary education).^23^

However, it would almost certainly be an overestimate to infer that such a decline directly translates into a reduction of the individual life span as suggested by the systematic socioeconomic differences in life expectancy in a society because only a part of those differences are in fact malleable. To ascertain YLL more precisely it is necessary to disentangle the determinants of the socioeconomic gap in life expectancy. Unfortunately, key determinants tend to overlap and only unfold their effects indirectly and in the long run, leaving life expectancy overdetermined. It is therefore difficult to specify the relative causal influence of fundamental sources such as the genetic disposition^24^ and SES^25,26^, the mechanisms through which they work such as health behaviors^27–29^ and morbidities (e.g. chronic diseases)^30,31^, and the immediate causes of death. Temporal and causal complexity as well as a lack of reliable data further complicate measurement immensely.^32,33^

Lately however, a number of studies made headway into ascertaining the individual contribution of factors such as income and education in the socioeconomic gap in life expectancy. In the European mean low income explains around 10-20% of an average 5-year gap in life expectancy between educational groups.^34^ For disability-adjusted life expectancy it is around 20% of a 8,5-year gap in life expectancy between educational groups.^35^ Educational and occupational status also account for around 20% of the 10-year gap in life expectancy between SES groups.^36,37^ In sum, income and educational status may each account for about a fifth (∼20%) of the socioeconomic gap in life expectancy.

The findings do not travel easily from the European high income countries to the rest of the world. In poorer countries morbidity and mortality are generally higher but health behaviors account for a smaller share of the socioeconomic differences in life expectancy.^31,38^ This plausibly leaves a larger share for socioeconomic factors. Education tends to entail a higher income premiums,^39^ but like health services is often not universally supplied and depends on personal income. The Socio-Demographic Index of the Global Disease Burden Project accounts for 85% of international differences in average healthy life expectancy by building the geometric mean of lagged per capita income, education of the population aged 15+, and the fertility rate of women aged 25+ (as a proxy for the standing of women in society).^40^ Against this background it seems plausible that in middle and low income countries, factors such as income and education may each account for 30% and more of the socioeconomic differences in life expectancy. In other words, the high, mid, and low income countries may have higher factors of socioeconomic determination of the life expectancy.

Based on these findings it is possible to construct a rough estimate of YLL due to permanent unemployment, reduction of working hours, and forgone education that entail a fall in SES group (from high to mid or mid to low). The YLL vary with the nominal socioeconomic gap in life expectancy (5y, 10y, 15y) and the degree of socioeconomic determination (SOD: 20%, 30%, 40%), depending on the overall level of income and development of a society. The two main components of the socioeconomic damage are due to income loss (YLLI) and forgone education (YLLE):

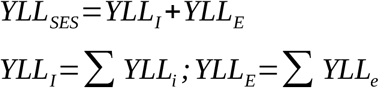

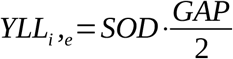

Educational loss in the pandemic further differentiates in two components: The most unfortunate cases where income loss or temporary school closures result in students permanently forgoing a higher educational bloc, depriving them of secondary or tertiary education (YLLe1); and the average income losses that affect the vast majority of students (YLLe2). Past examples show that even short episodes of temporary school closures have a measurable average impact on income in later life. The first pandemic-related school closures may reduce lifetime earnings by 1-4%, depending on the subsequent ability for learning compensation.^41,42^ Adding the second phase of school closures at turn of the year 2020/21 and considering that longer closures add exponentially, current losses in lifetime earnings may amount to 5% on average (or about one eights of a decline in SES group). For many low to middle income countries this is possibly an underestimate, given that school closures were on average longer, potentially entail higher dropout rates, and would have held a higher income premium.^43,44^

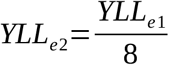

### 2.1 Covid-YLL

Attributing YLL to Covid-19 is no less complicated and uncertain than attributing YLL to the socioeconomic consequences of the pandemic. Various studies on high income countries that draw on aspirational national life tables estimate the loss of life due to Covid-19 at 10-13 YLL per death.^45,46^ Accounting for co-morbidities results in a reduction by about 1-3 YLL.^47^ A different approach compares the age distribution of Covid-19 death with the age distribution for all cause mortality in a birth cohort. The approach, that likely is an underestimate of the remaining life years of an individual, estimates the Covid-19 toll to be around 2-5 YLL per deaths.^48,49^ A third approach, using data on care home mortality and average lengths of stay, arrives at an estimate of 5 YLL for the age bracket 70+.^50^ Complementing the latter approach with YLL estimates from life tables for under 70-year-olds would leave the overall societal average at around 7-9 YLL, depending on whether the group accounts for 10% or 15% of Covid deaths.

Amid these uncertainties, the article suggests correcting the aspirational YLL from the life tables for socioeconomic differences in life expectancy. In other words, to drop the assumption of a poverty-free and egalitarian society. This may improve YLL estimates in two ways: a more accurate attribution of YLL to its fundamental and immediate causes; and a potentially more precise estimate of the actual YLL of an individual without giving up on the normative claim of not accepting a lower than ideal life expectancy. To that end, country-specific findings on socioeconomic differences in life expectancy should be combined with data on seroprevalence, hospitalization rates, and deaths among groups with a low SES compared to the general population.

Studies consistently find that people with a low SES are significantly more affected than people with a high SES. In high income countries around half of all Covid-19 deaths occur among people with a low SES. In the US, people with below median income account for two thirds of deaths (the lowest tertile for about half).^51^ The poorest quintile suffers from a third more co-morbidities and twice the case count and death rate.^52^ In Scotland, the lowest quintile may account for half of all ICU admissions.^53^ Also, in the otherwise more equitable Germany and Sweden people with low SES account for 40-60% of hospitalizations and death.^54,55^

Undertaking similar estimate for low and middle income countries faces severe data challenges. The only large cross-country comparison to date, using national UN WPP life tables for remaining life expectancy at the exact age of deaths, estimates mean YLL per capita at 13 YLL for high income countries and 19 YLL for middle and low income countries.^56^ Their estimates are used in the following examples. The profile of Covid deaths in low and middle income countries is more ambiguous. Similar to all-cause mortality Covid deaths tend to be younger. While this reduces case mortality, high levels of inequality and poverty come with additional risks for the younger. In other words, fewer people die but those that die lose more life years. Socioeconomic factors associated with a low SES such as poor living conditions and work in the informal economy, which often accounts for 50-90% of employment, seem to play a key role in the spread and higher mortality in the younger age brackets.^57–59^ It is therefore reasonable to assume that the share of Covid deaths with a low SES is even higher in low and middle income countries.

To give an example, in high income countries with moderate socioeconomic inequalities, a 7,5-year socioeconomic gap in life expectancy, and 40% of Covid-19 deaths in the group with low SES, 4.5 YLL can be attributed to the SES and would have to be subtracted from the nominal YLL from the life tables. In the case of Sweden (10.3) or Germany (10.6) that would leave 5.8 and 6.1 YLL, respectively, as attributable to Covid-19. In more unequal countries with a socioeconomic gap in life expectancy of 10-years, and 50% of Covid deaths among people with a low SES, 6.5 YLL would be subtracted from the nominal YLL. For the United States (14.8) or Brazil (16.4), for example, this would leave 8.3-9.9 YLL attributable to Covid. For lower income countries with high levels of poverty and large lifespan inequalities, the aspirational 19 YLL from the life tables may have to be reduced even more by 10-12 YLL to 7-9 YLL due to Covid. In low income countries the socioeconomic gap in life expectancy looks somewhat different and entails higher life span inequality in general, probably because absolute and relative poverty overlap. In sum, when accounting for socioeconomic differences in life expectancy YLL attributable to Covid may be between 6 and 10 YLL, depending on levels of income and inequality in a society. In other words, around half of YLL from the national life tables may in fact be socioeconomically determined. The global average for years of life lost per Covid deaths (∅ YLLcov) may therefore be at around 8 years with somewhat lower values for more egalitarian high income countries (∼6 YLL) and somewhat higher values for more unequal and poor countries (∼10 YLL).

Current empirical projections for the global pandemic estimate up to 5 million deaths by August 2021.^1^ With vaccine role out slow in most parts of the world and uncertain protection against new virus variants, it cannot be ruled out that this number multiplies over the next years. For the following calculations six hypothetical scenarios (W1-6) with different number of Covid deaths are constructed.

The loss of life years in these scenarios is then juxtaposed with the socioeconomic damage in the pandemic to ascertain proportionality. The main idea is to calculate the number of people affected by the socioeconomic damage that would entail the equivalent amount of YLL.

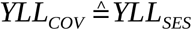

To that end, the individual components of socioeconomic damage YLLses are distributed among subgroups of the globally affected population (Ng): students (ne), workers (ni), and people living in extreme poverty (np).

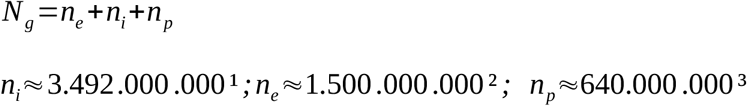

1 ILO; 2 UNESCO; 3. World Bank.

Learning loss is divided into two subgroups. Those students suffering the average learning and subsequent income loss from school closures YLLe2 and the worst hit students that forgo a 3-4 year higher learning block YLLe1 (i.e. additional dropouts due lack of funding or qualification for higher education). Because students with a high SES may have more capacities to compensate for learning losses it is assumed that two thirds of the students worldwide (=0.9 billion) suffer the average learning and subsequent income loss due to school closures.

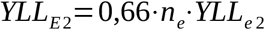

The resulting value is subtracted from the overall years of life lost due to Covid (YLLcov). The remaining damage is then distributed among the students with a learning block loss (YLLE1), people with an income loss YLLI, and those that fall into extreme poverty as a result YLLP.

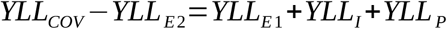

Each group carries a weighted burden that reflects group size and the social gradient (α,β,γ). Income losses account for slightly more than half (0.54) and forgone education (0.23) and poverty (0.22) each for slightly less than a quarter of all YLLs.

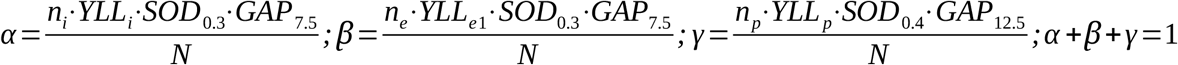

With these shares it is possible to individually calculate the total number of workers (Xi), students (Xe), and poor (Xp) for whom the socioeconomic damage in the pandemic would have to become *permanent*.

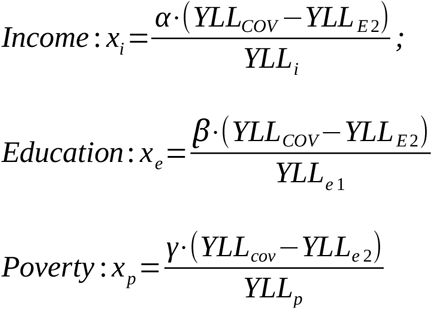

## 3. Results

The standard model reflects specifications for the global average (8 YLL per Covid death, a 7,5-year socioeconomic gap in life expectancy, and a SOD of 0.3). The tables read as follows. Each row provides the total number of workers, poor, and students for which the socioeconomic damage would have to become *permanent* for the YLL to be equivalent of those attributable to Covid-19. Lines are gray for negative values, i.e. when the average socioeconomic damage from school closures is higher than the YLL due to Covid. A separate column provides the common percentage share, which by definition is identical for all groups.

In the first three pandemic scenarios with up to 10 million Covid deaths school closures alone amount to more YLL. While that does not automatically make them disproportionate, it raises the burden of proof for their necessity and efficacy and illustrates the urgent need to compensate for learning loss. Only the subsequent scenarios W4-6 would justify single-digit percentage increases in the share workers with income losses due to unemployment and reduction of working hours (∼48.7-203.3 million), people that fall into extreme poverty (9-37.5 million), and students that drop out of current education or do not qualify for a higher learning block (20.9-87.3 million).

Because at the same time the average per person damage to life from temporary school closures is comparatively minor and may be deemed acceptable by society, there is a case for excluding it from the analysis. Dropping YLLE2 from the calculation results in an increase of the percentage shares in the previously disproportionate scenarios W1-3 to 0.6-1.1%. The absolute numbers for proportional socioeconomic damage thus remain relatively small in the first three scenarios. That illustrates the potential harm to life from only comparatively minor socioeconomic consequences. Subsequent scenarios roughly represent a shift by one scenario compared to the model including school closures. The percentage shares in W4-6 increase from 1.4-5.8% to 3.3-7.7%. In the worst case scenario 270 million people suffer from income loss, 50 million fall into extreme poverty, and 116 million forgo a higher learning block.

The results highlight the urgent need for compensating existing and avoiding further socioeconomic damage in the pandemic. Avoiding a relatively minor number of 5 million people with income loss, 1 million additional people in poverty, and 2 million with a higher learning loss saves a similar amount of life years as saving more than 1 million people from dying from Covid-19. Current predictions of the long-term socioeconomic consequences in the pandemic are uncertain but illustrate the effort that would be required. According to the ILO, the equivalent to 255 million full time jobs was lost in 2020, including around 114 full time jobs and two to three hundred million working hour reductions.^2^ Depending on which scenario becomes reality over the next years, 60-90% of those job losses would have to be saved from becoming permanent for the loss in life years not to exceed the YLL due to the pandemic. Looking at educational loss one year into the pandemic, UNESCO estimates that more than half of all school children continue to face learning disruptions and 24 million (1.6%) students are at risk of dropping out completely, not including those that will not qualify for higher education.^60^ Educational loss is thus similar to levels estimated for scenario W4. Furthermore, around 100-150 million people fell into extreme poverty due to the pandemic.^4,5^ The number almost doubles to 228 million for the higher poverty threshold of $3.20 instead of $1.90 per day. Even in the worst case scenario these losses must not become permanent for more than 20-40% of those that recently fell into (extreme) poverty.

For a second set of results the standard model specifications are adapted to reflect different country conditions (see Table 6). The three columns on the left assume conditions more similar to a typical high income country. The SOD is at 20%, the average per capital YLL due to Covid are at 6 YLL, and the socioeconomic gap in life expectancy varies between 5 YLL (South Europe), 7,5 YLL (Central, Western Europe) and 10 YLL (Eastern Europe, US). The middle columns show two variations of the standard specification (marked red) with a higher socioeconomic gap in life expectancy (10 and 12.5 years). The three columns on the right assume conditions more characteristic for low income countries with a high level of socioeconomic determination (40%), a high loss of per capita life years due to Covid (10 YLL), and a wide socioeconomic gap in life expectancy (10-15 years). For reasons of readability the table presents the changing values only as the common percentage share. In theory it is possible to infer from the percentage values to the total numbers for each group – a 1% change corresponds to ∼35 million people with income loss, 15 million students with forgone higher education, and 0.64 million people falling into extreme poverty. However, those absolute values are of limited empirical insight because the model merely assumes that whole world behaves like the respective income group.

**Table 1:** YLLi,e (per capita) for decline in SES.

| SOD \ GAP |  | 5 | 10 | 15 |
| --- | --- | --- | --- | --- |
| High income | 0,2 | 0,5 | 1 | 1,5 |
| Middle income | 0,3 | 0,75 | 1,5 | 2,25 |
| Low income | 0,4 | 1 | 2 | 3 |

**Table 2:** YLLs due to SES.

| **Covid death**<br>**Per SES groups**<br>**(low/mid/high) (in %)** \ **GAP** | **5** | **7.5** | **10** | **12.5** | **15** |
| --- | --- | --- | --- | --- | --- |
| **40/40/20** | 3 | 4.5 | 6 | 7.5 | 9 |
| **50/30/20** | 3.3 | 4.9 | 6.5 | 8.1 | 9.8 |
| **60/30/10** | 3.8 | 5.6 | 7.5 | 9.4 | 11.3 |
| **70/20/10** | 4 | 6 | 8 | 10 | 12 |

**Table 3:** Scenario of Covid-19 deaths worldwide.

| Scenario | Covid deaths | YLL(cov)<br>(Ø6) | YLL(cov)<br>(Ø8) | YLL(cov)<br>(Ø10) |
| --- | --- | --- | --- | --- |
| W1 | 5 Mio. | 30 Mio. | 40 Mio. | 50 Mio. |
| W2 | 7,5 Mio. | 45 Mio. | 60 Mio. | 75 Mio. |
| W3 | 10 Mio. | 60 Mio. | 80 Mio. | 100 Mio. |
| W4 | 30 Mio. | 180 Mio. | 240 Mio. | 300 Mio. |
| W5 | 50 Mio. | 300 Mio. | 400 Mio. | 500 Mio. |
| W6 | 70 Mio. | 420 Mio. | 560 Mio. | 700 Mio. |

**Table 4:** Equivalent socioeconomic damage (standard model)

| Scenario | Covid deaths | Income loss<br>(Total) | Covid poor<br>(Total) | Education<br>loss<br>(Total) | Share<br>(in %) |
| --- | --- | --- | --- | --- | --- |
| **W1** | **5 Mio.** | -47,9 Mio. | -8,8 Mio. | -20,6 Mio. | -1,4% |
| **W2** | **7,5 Mio.** | -38,3 Mio. | -7,1 Mio. | -16,4 Mio. | -1,1% |
| **W3** | **10 Mio.** | -28,6 Mio. | -5,3 Mio. | -12,3 Mio. | -0,8% |
| **W4** | **30 Mio.** | 48,7 Mio. | 9 Mio. | 20,9 Mio. | 1,4% |
| **W5** | **50 Mio.** | 126 Mio. | 23,2 Mio. | 54,1 Mio. | 3,6% |
| **W6** | **70 Mio.** | 203,3 Mio. | 37,5 Mio. | 87,3 Mio. | 5,8% |

**Table 5:** Equivalent socioeconomic damage (excluding school closures, YLL(E2))

| **Scenario** | **Covid deaths** | **Income loss**<br>**(Total)** | **Covid poor**<br>**(Total)** | **Education**<br>**loss**<br>**(Total)** | **Share**<br>**(in %)** |
| --- | --- | --- | --- | --- | --- |
| **W1** | **5 Mio.** | 19,3 Mio. | 3,6 Mio. | 8,3 Mio. | 0,6% |
| **W2** | **7,5 Mio.** | 29 Mio. | 5,3 Mio. | 12,5 Mio. | 0,8% |
| **W3** | **10 Mio.** | 38,7 Mio. | 7,1 Mio. | 16,6 Mio. | 1,1% |
| **W4** | **30 Mio.** | 116 Mio. | 21,4 Mio. | 49,8 Mio. | 3,3% |
| **W5** | **50 Mio.** | 193,3 Mio. | 35,6 Mio. | 83 Mio. | 5,5% |
| **W6** | **70 Mio.** | 270,6 Mio. | 49,9 Mio. | 116,2 Mio. | 7,7% |

**Table 6:** Proportional socioeconomic damage across different model specification.

|  | SOD | 0,2 |  |  | 0,3 |  |  | 0,4 |  |  |
| --- | --- | --- | --- | --- | --- | --- | --- | --- | --- | --- |
|  | Ø YLL | Ø 6 |  |  | Ø 8 |  |  | Ø 10 |  |  |
|  | GAP | 5 | 7,5 | 10 | 7,5 | 10 | 12,5 | 10 | 12,5 | 15 |
| <=0% | W1 | -3 % | -2 % | -2 % | -1 % | -1 % | -1 % | -1 % | -1 % | 0 % |
| 0-<1% | W2 | -3 % | -2 % | -1 % | -1 % | -1 % | -1 % | 0 % | 0 % | 0 % |
| 1-2,9 | W3 | -2 % | -2 % | -1 % | -1 % | -1 % | 0 % | 0 % | 0 % | 0 % |
| 3-4,9 | W4 | 1,3 % | 0,8 % | 0,6 % | 1,4 % | 1 % | 0,8 % | 1,3 % | 1 % | 0,8 % |
| 5-9,9% | W5 | 5 % | 3,3 % | 2,5 % | 3,6 % | 2,7 % | 2,2 % | 2,8 % | 2,2 % | 1,9 % |
| >10% | W6 | 8,7 % | 5,8 % | 4,4 % | 5,8 % | 4,4 % | 3,5 % | 4,4 % | 3,5 % | 2,9 % |

The different model specifications obtain two main results. One at the cross-country level and one at the within-country level. First, the differences in parameters tend to largely even out across the different model specifications. The first three scenarios remain disproportionate in low, middle, and high income countries. The results also largely hold when dropping the average damage from school closures (YLLE2) (Table 7). Larger differences only occur in scenario W4-6. At the extreme ends of the model specifications, the proportionality of the socioeconomic damage differs by the factor 3 (2,9-8,7%), reflecting the steeper social gradient in low income countries. The second main result is that at constant YLLs per Covid death the socioeconomic damage becomes disproportionate much earlier in more unequal societies. In more egalitarian high income countries twice the socioeconomic damage is proportional than in the most unequal ones. In middle and low income these differences are less pronounced but remain significant.

**Table 7:** Proportional socioeconomic damage across different model specification (excluding YLLe2)

|  | SOD | 0,2 |  |  | 0,3 |  |  | 0,4 |  |  |
| --- | --- | --- | --- | --- | --- | --- | --- | --- | --- | --- |
|  | Ø YLL | Ø6 |  |  | Ø8 |  |  | Ø10 |  |  |
|  | GAP | 5 | 7,5 | 10 | 7,5 | 10 | 12,5 | 10 | 12,5 | 15 |
| <=0% | W1 | 1 % | 1 % | 0 % | 1 % | 0 % | 0 % | 0 % | 0 % | 0 % |
| 0-<1% | W2 | 1 % | 1 % | 1 % | 1 % | 1 % | 0 % | 1 % | 0 % | 0 % |
| 1-2,9 | W3 | 2 % | 1 % | 1 % | 1 % | 1 % | 1 % | 1 % | 1 % | 1 % |
| 3-4,9 | W4 | 5,6 % | 3,7 % | 2,8 % | 3,3 % | 2 % | 2,0 % | 2,3 % | 2 % | 1,6 % |
| 5-9,9% | W5 | 9 % | 6,2 % | 4,7 % | 5,5 % | 4,2 % | 3,3 % | 3,9 % | 3,1 % | 2,6 % |
| >10% | W6 | 13,1 % | 8,7 % | 6,5 % | 7,7 % | 5,8 % | 4,6 % | 5,4 % | 4,4 % | 3,6 % |

## 4. Discussion

The article set out to narrow in on the difficult question of proportionality between the health and socioeconomic damage in the pandemic. YLL were suggested as a common measure of comparison for the two dimensions. Correctly attributing YLL to Covid-19 and SES, requires disentangling the fundamental and immediate sources of life expectancy. In order to use the life table approach that is common in YLL estimates, the article made an attempt at dropping the inherent assumption of a poverty-free and egalitarian world (or society). The discussion concluded that up to half of the average per capita YLL from the life tables may in fact be socioeconomically determined. Because SES is associated with morbidity and mortality which in turn is similar for Covid-19 and all-cause mortality, the approach may yield analytic benefits beyond the current pandemic.^61,62^ Ecological data on the SES of the population of interest may proxy for a lack of individual-level data on the prevalence of morbidity and other risk factors.

The application to the pandemic highlights the difficult trade-offs involved in the short- and long-term protection of health. While NPIs target immediate health concerns, the long-term socioeconomic damage is likely to entail a steep cost to life that requires immediate attention in the aftermath of the pandemic. Avoiding 3 million additional cases of extreme poverty may come with a similar life tag as protecting 1 million people from dying from Covid-19. In countries that lack the necessary resources to compensate for the socioeconomic damage in the pandemic, more drastic NPIs such as business and school closures need to be considered very carefully.^7^ If not compensated for, school closures are likely to be disproportionate even in high income countries. At the very least they carry a significant burden of proof regarding their suitability and necessity.

Interestingly, the question of proportionality has otherwise been rather similar across different income groups, largely because the social gradient and the associated loss of life is steeper for both Covid-19 and the NPIs. Sill, levels of within-country inequalities may be a key concern regarding the proportionality of the NPIs. This is especially true because a wider socioeconomic gap in life expectancy signals a weaker social safety net that could compensate for losses.

The approach comes with a number of important limitations. Even though measurement issues are likely to outweigh causal concerns by far, the assumptions on the extent of socioeconomic determination of the life expectancy and the size of the socioeconomic gap in life expectancy in low and middle income countries is based on rather weak empirical evidence. Furthermore, the model does not account for the non-lethal health impact, for example, from “Long-Covid” or the psychosocial consequences from NPIs. Future research could inform an extended estimate using quality-adjusted estimates such as the healthy life expectancy (HALE). Finally, the model does not account for the Covid-related disease burden on economic activity which is sometimes invoked to justify harsher NPIs as in fact reducing the socioeconomic damage in the pandemic. However, existing research into the relationship has thus far been unable to disentangle the causal role of voluntary behavioral change, formal and informal NPIs, and the objective disease burden in reducing economic activity.^63^

## Data Availability

Data openly available in a public repository that does not issue DOIs.

